# Social, environmental and economic risk factors and determinants of sleep disturbances in Latin America: A systematic review and a meta-analysis of public health literature

**DOI:** 10.1101/2023.06.02.23290915

**Authors:** Faustin Armel Etindele Sosso, Filipa Torres Silva, Rita Queiroz Rodrigues, Margarida Carvalho, Sofia Zoukal, Gabriel Cordova Zarate

## Abstract

**Background:** Mental health recovery is associated with healthy sleep, and disturbances of both, areas represent an increasing public health issue worldwide, particularly in Latin America. Among multiple determinants affecting sleep health, individual’s socioeconomic status (SES) is the most ignored and underestimated through literature. No systematic review on the relation between SES and sleep health has been previously conducted in Latin America.

**Methods:** PRISMA guidelines were used.

**Results:** Twenty cross-sectional studies were selected. 80,0% (n=16) were performed in Brazil, 10,0% (n=2) in Peru, 5,0% (n=1) in Chile, and 5,0% (n=1) were multicentric (11 countries). The combined total number of participants were N=128455, being 3,7% (n= 4693) children, 16,0% (n= 20586) adolescents and 80,3% (n=103176) adults. Higher SES was associated with lower sleep duration. Lower SES was associated with a decrease in sleep quality. Excessive Daytime Sleepiness (EDS) was significantly more prevalent in individuals with lower family income and less education. Sleep bruxism was associated with higher education and lower SES was associated with more sleep bruxism. A meta-analysis of random effects model showed that pooled prevalence of sleep disturbances was 27.32 % (95 % CI 21.71–33.75) with high heterogeneity (I^2^ = 100 %). Pooled prevalence decreased with high education (OR 0.83; 95%CI [0.75-0.91]; I^2^= 79%), while it increased by low income (OR 1.26; 95%CI [1.12-1.42]; I^2^= 59%), unemployment (OR 2.84; 95%CI [2.14-3.76]; I^2^= 0%) or being housewife (OR 1.72; 95%CI [1.19-2.48]; I^2^=55.4%).

**Discussion:** Gradient of health disparity existing for some diseases like cardiovascular illness, seems the same for sleep disturbances regardless of world region. Therefore, sleep disturbances management should be address in a multidimensional approach with a significant investment of government in targeted public health program, to reduce sleep disparities and support research before the situation become uncontrollable.

## 1 INTRODUCTION

Promoting recovery in mental health is an ongoing project which never ends, because mental health is a complex public health outcome requiring multidimensional interventions at the economic level, the populational level and societal level [1,2]. Mental health is also highly dependent of sleep which has a recognized impact on several brains functions [3] and global health status as well as stress [4]. In addition, sleep has a significant influence on mental health due to its relationship with people’s socioeconomic status (SES) and its connection with multiple biological systems involved in neurological disorders such as anxiety [1,5–10]. That means in others words that, sleep disturbances are mental disorders resulting from complex socioecological and economic interactions between brain, society where we live, global health and SES [7,11,12]. Thus, sleep health inequalities represents a mental health outcome similar to public health issues previously reported for cardiovascular, mental health and metabolic diseases [11]. Empirical literature in western countries seems validated the hypothesis stating that, low SES individuals reported more sleep disturbances than high SES people [13–18]. Similar evidence exists in Asia [19,20] and Oceania [21,22]. Thus, sleep health disparity could be public health issues in other regions, such as Africa and Latin America. Few studies documented sleep health disparities in South and Central America due to a wide range of determinants such as employment, income, education, occupation and social position [11,23–25]. A comprehensive evaluation of public health literature related to Latin America showed that no previous systematic review has investigated the relationship between SES and sleep health in this region. It’s important to analyze if trends related to the influence of SES determinants of mental health on sleep health observed in western countries are following the same patterns in Latin America. The goals of this systematic review are to 1) document the socioeconomic determinants of sleep health in Latin America and 2) promote the mental health recovery related to a good sleep health.

## 2 METHODS

### 2.1 Literature search

Relevant peer review studies included in this systematic review were identified by searching the following databases: Web of Science, Scopus, PubMed/Medline and Ovid, restricted on a period between January 1990 and May 2023. A combination of terms “socioeconomic”, “socio-economic”, ‘’social position’’, ‘’social class’’, ‘’socioeconomic position’’, “determinant*”, “indicators*”, “markers*”, “health inequalities”, “health inequities”, “sleep”, ‘’sleep disorders’’, ‘’sleep disturbances’’, ‘’sleep complains’’, “sleep outcome”, “sleep health”, “south america*”, “southern america*”, “central america*”, “latin america” was used. All included articles were identified following the PRISMA guidelines detailed in Figure 1 [25].

**Figure 1.**
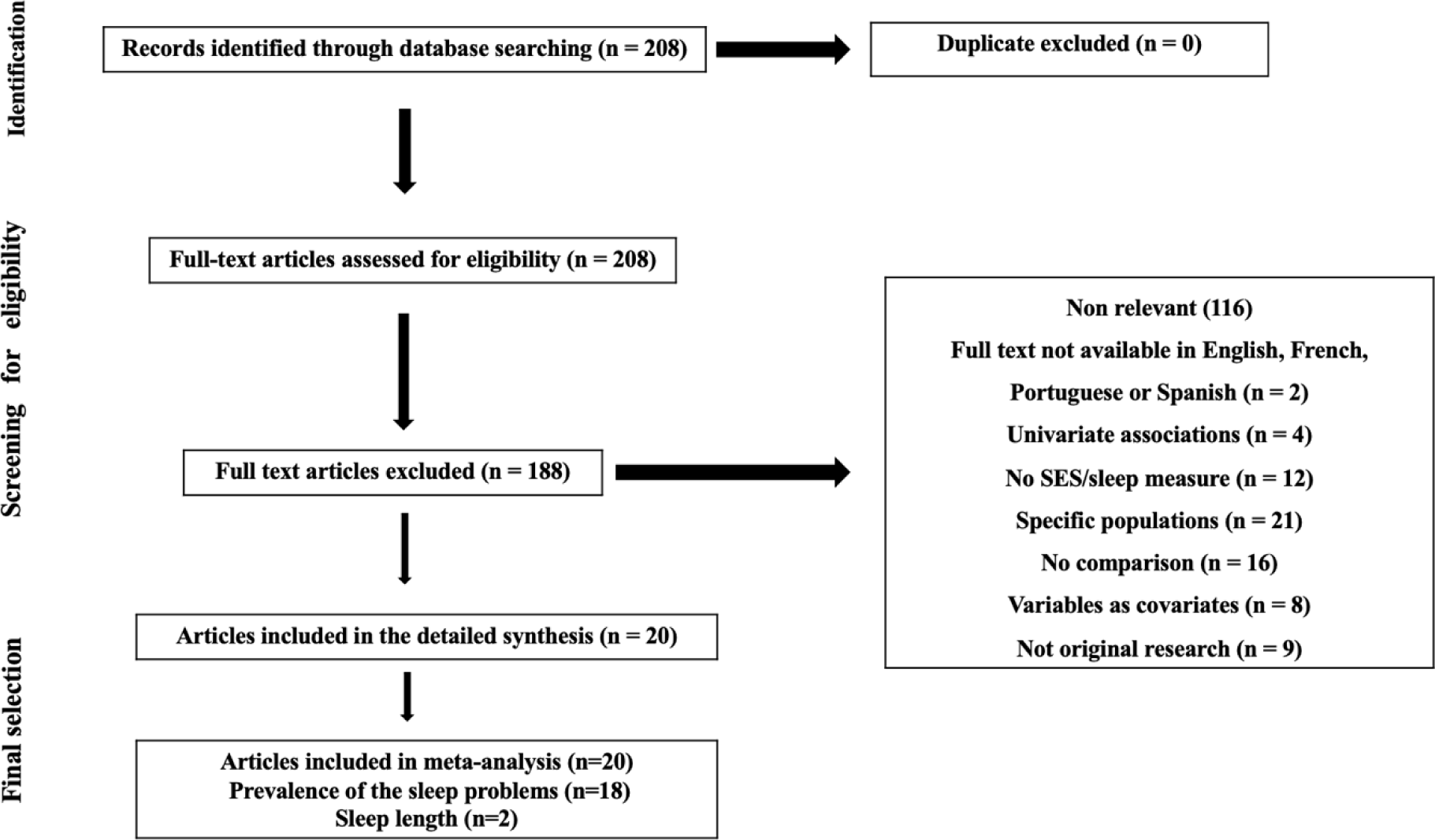
PRISMA flowchart of study selection process

### 2.2 Selection of evidence and data extraction

Two reviewers (*FAES, SZ*) independently reviewed the titles and abstracts of the studies identified by the search strategy and determined eligibility for inclusion, while disagreements were resolved by consensus with a third external reviewer. For studies that passed the initial screening, the entire text was collected and the screening process was repeated by the same co-authors to conclude with the final articles included, validated by the third reviewer. Then, four reviewers (FTS, RQR, MC, FAES) extracted from each report the following studies characteristics: population, % women, age, sample size, SES measures, relevant statistics, interaction or mediation, sleep measures, conclusions/main effects, statistical methods and results significance.

### 2.3 Inclusion and exclusion criteria

Empirical studies were defined as studies of any design (cross-sectional, retrospective or longitudinal) assessing humans of any gender, race/ethnicity and age from the general population of any country from Latin America. The article should include an objective or a subjective measure of SES, such as income, educational level, wealth, profession/occupation, employment status, perceived SES or self-reported SES. Proxy measures of SES, such as neighborhood and social class, were also included. For studies involving children and adolescents, perceived family SES measures, such as parental education, parental occupation or annual household income were utilized. Studies were excluded based on the following criteria: 1) They were interventional trials, any type of reviews (narrative, overview, systematic, umbrella, meta-analyses), case series, case reports, conference series or any writing without original research (editorial, commentary, letter to editors), 2) articles did not provide statistical significance in cases where either SES or sleep were evaluated as covariates or mediators, 3) the full text was not accessible, 4) authors/researchers recruited participants with medical conditions at the beginning of study (for example people with medications including sleep pills, people with cancers, people with neurodegenerative diseases, etc.…) and 5) articles were not written in English, French, Portuguese and Spanish (languages of authors).

### 2.4 Quality rating of studies

*The National Institute of Health’s (NIH) Quality Assessment Tool for Observational Cohort and Cross-Sectional Studies* was used to rate the quality of included studies [26]. This quality rating’s tool analyzed fourteen quality criteria asking an equal number of questions about study objectives, population, exposures, outcomes, follow-up rates, and statistical analysis. SES was considered the exposure variable and sleep measures were the outcome variable, respectively. Overall quality ratings (**Table 2**) were calculated by taking the proportion of positive rating for the sum of applicable criteria. Studies with < 50% positive rating were judged as poor quality, ≥ 65% as good quality, and the rest as fair quality [26].

### 2.5 Study outcomes

The primary outcomes were related to the prevalence of sleep disturbances in Latin America and the difference in sleep length between men and women. The secondary outcomes aimed to explore how SES influences the prevalence of sleep disturbance in Latin America. This exploration was conducted through analyzing education, income, employment status, and perceived SES, as long as they were reported by at least two independent studies.

### 2.6 Data analysis

Meta-analysis was performed using the meta package on R with RStudio interface (Version 4.1.3, R Core Team, 2022) to analyze the collected data.

In each study, the prevalence of sleeping disturbances in Latin America was obtained. Studies did not report the prevalence are excluded. The random effects model was used with the logit transformation for obtaining the pooled results, because it produces more conservative results than fixed-effects models regardless of heterogenicity scores [27]. The pooled prevalence estimates of sleeping disturbances in Latin America was presented as a percentage with 95% confidence intervals (CIs) using a forest plot. Also, a subgroup analysis was conducted for different sleep issues, cities, study’s quality and age group to assess the contribution of each study to overall heterogeneity in the prevalence of sleep disturbance.

To measure the relationship between SES and sleeping disturbances, we extracted from the selected publications the adjusted odds ratio (aOR) with 95% CIs. Their standard errors were calculated from the respective CIs. The value from each study and the corresponding standard error were transformed into their natural logarithms to stabilize the variances and to normalize the distribution. The pooled OR (and 95%CI) was estimated using a DeirSimonian-Laird random effect model. In situations in which a study reported effect estimates for independent subgroups, the subgroups were treated as individual studies in the meta-analysis.

The test of the overall effect was assessed by using Z-statistics at p<0.05. The heterogeneity among the included studies was assessed using Cochran’s Q test and I^2^ statistics. The thresholds of 25%, 50%, and 75% were used to indicate low, moderate, and high heterogeneity, respectively [28, 29]. A funnel plot based on Egger’s regression test was used to evaluate the publication bias [30]. In all analysis, a *p*-value less than 0.05 was considered statistically significant.

## 3 RESULTS

### 3.1 Characteristics of included articles

Twenty articles were included [31–50] in the final sample, all cross-sectional studies and available in **Table 1**. Among these studies, 80,0% (n=16) were performed in Brazil [31–33, 35, 37, 38, 41–50], 10,0% (n=2) in Peru [39, 40], 5,0% (n=1) in Chile [34], and 5,0% (n=1) were multicentric (11 countries)[36]. The combined total number of participants were N=128455, being 3,7% (n= 4693) children, 16,0% (n= 20586) adolescents [38, 39, 41, 42, 45] and 80,3% (n=103176) adults [31–37, 39, 40, 46–50]. The smallest sample size was n=851 [43] and the largest was n=60202 [49]. The socioeconomic indicators used were income (monthly personal income, monthly family income, per capita income, annual household income) [31–35, 37, 38, 41, 43–47], wealth/assets [37, 39, 40, 49, 50], number of residents in the household [46], employment status/occupation [31, 32, 34, 35, 37–39, 46, 47, 50], accessed healthcare system [37] and composite indices [42]. The sleep variables used were excessive daytime sleepiness [32, 40, 49], sleep duration [37–39, 41, 44–46, 50], sleep quality/sleep disturbance [33, 34, 36, 38, 44, 47, 49], insomnia [31, 36], obstructive sleep apnea (OSA)/sleep-disordered breathing (SDB) symptoms [35, 40] and bruxism [42, 43, 48]. Majority of articles were of poor quality (55%) and detailed qualitative evaluation is available in **Table 2**.

**Table 1.**
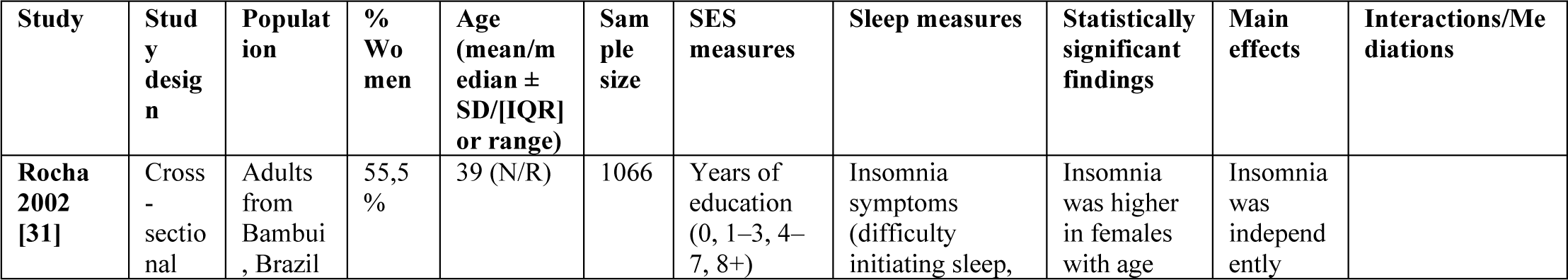
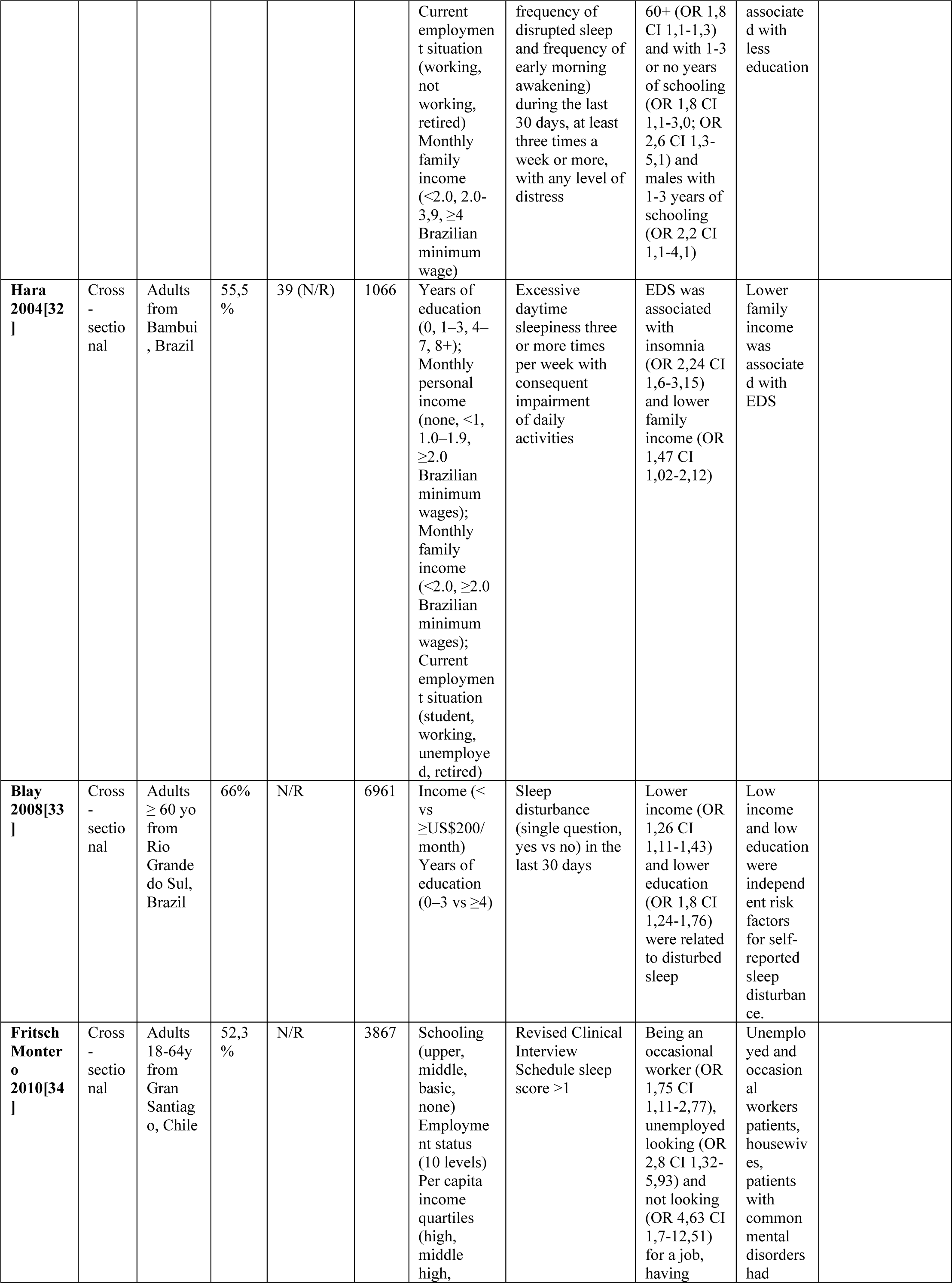
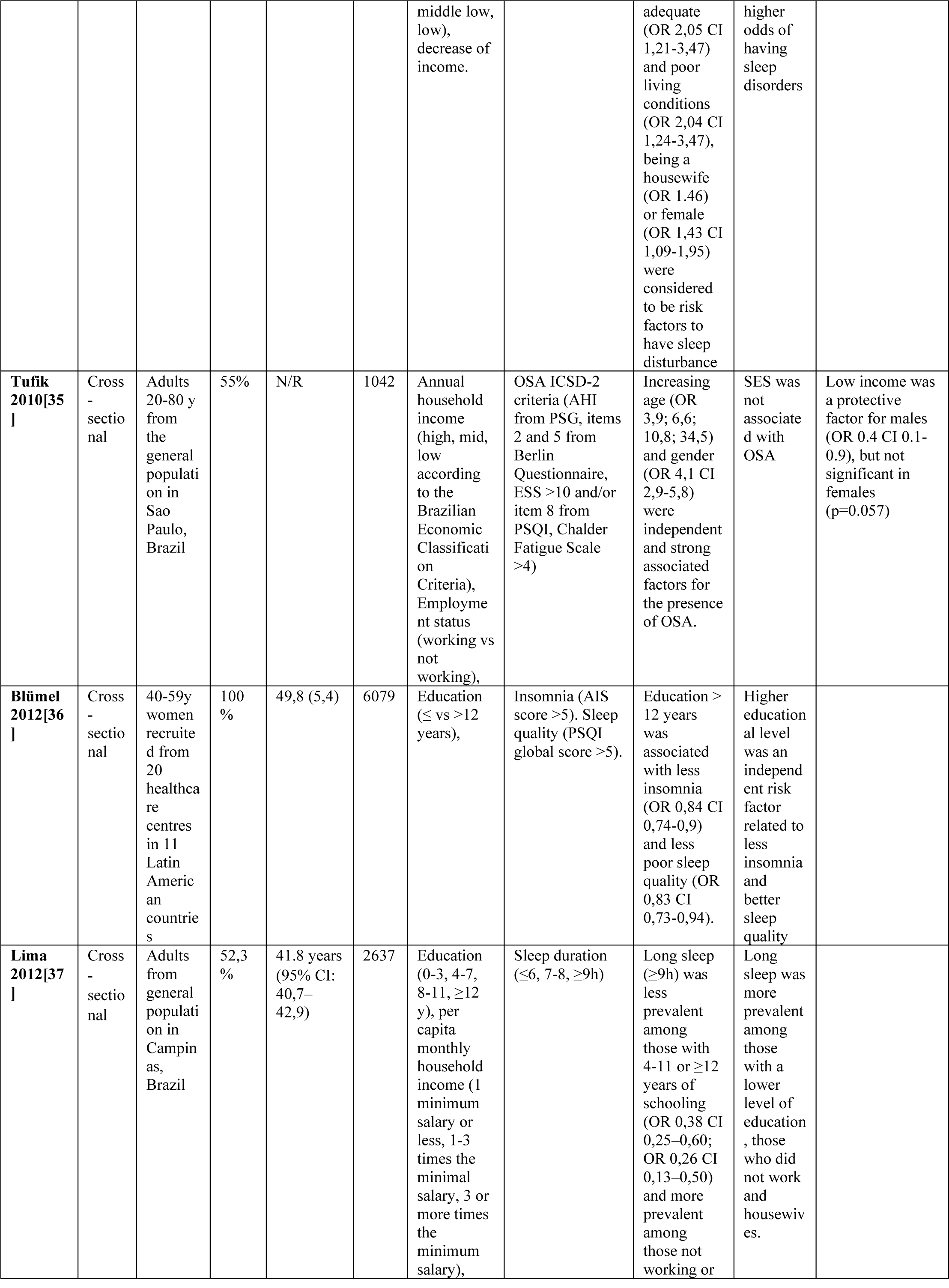
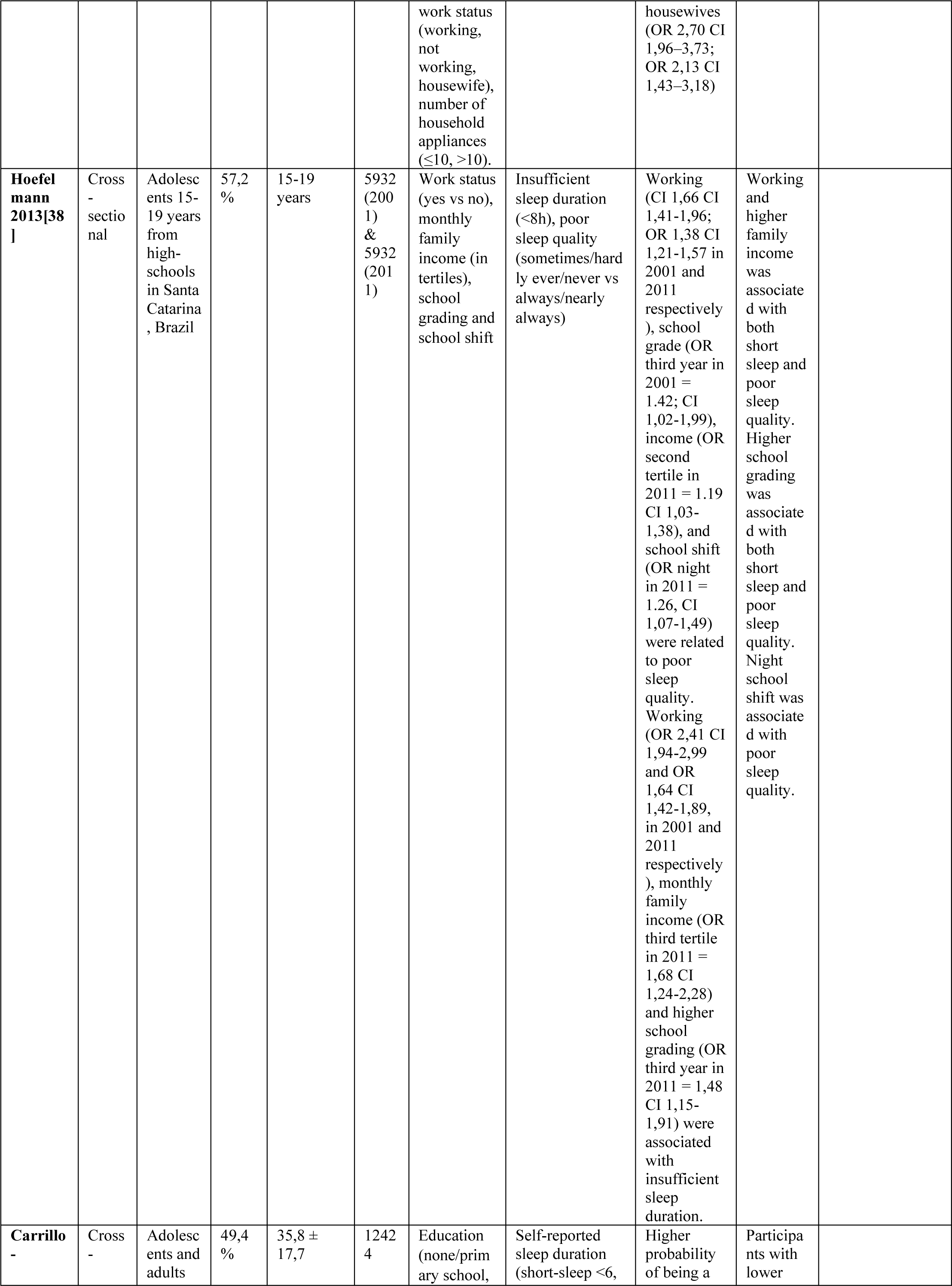
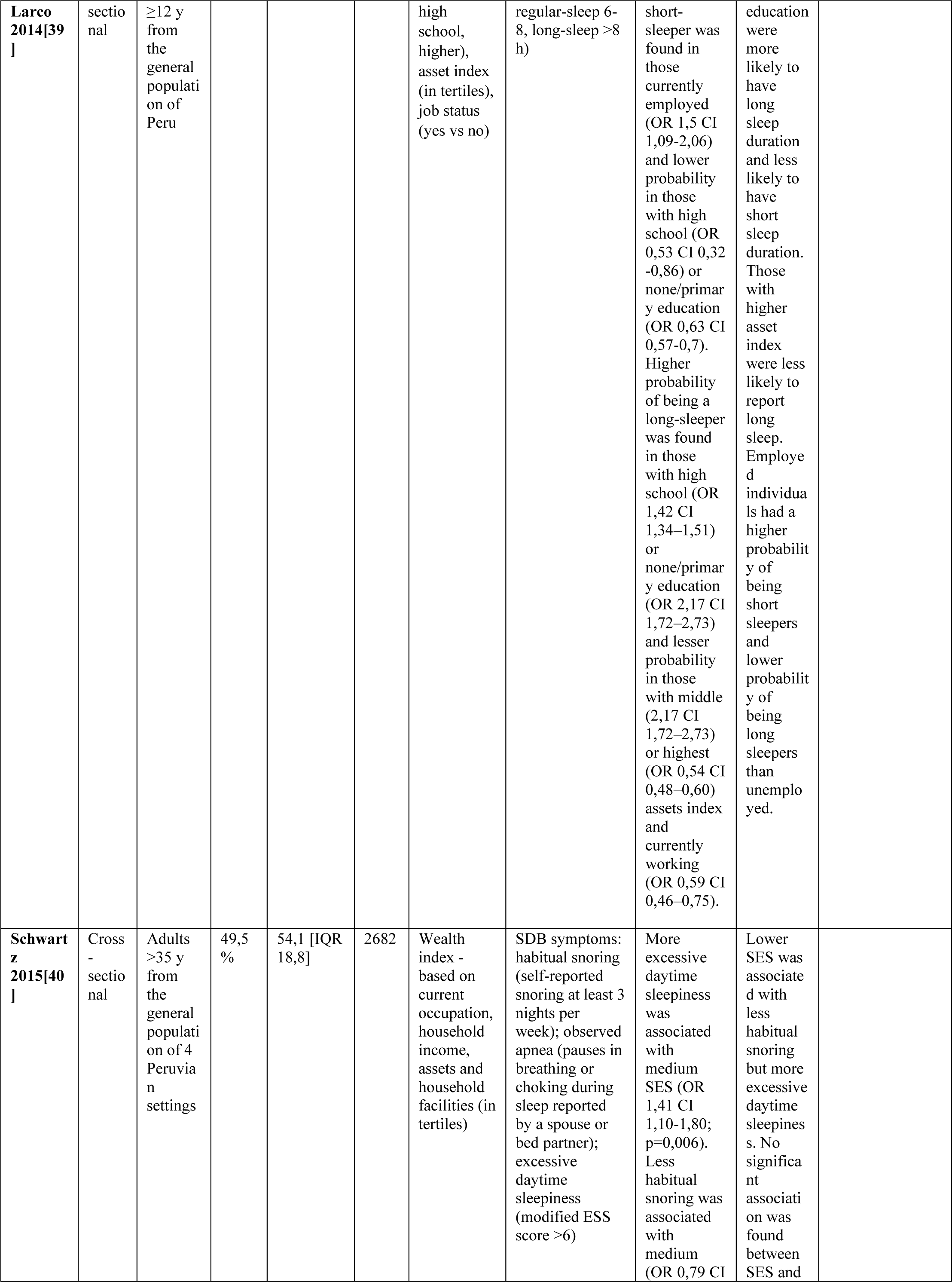
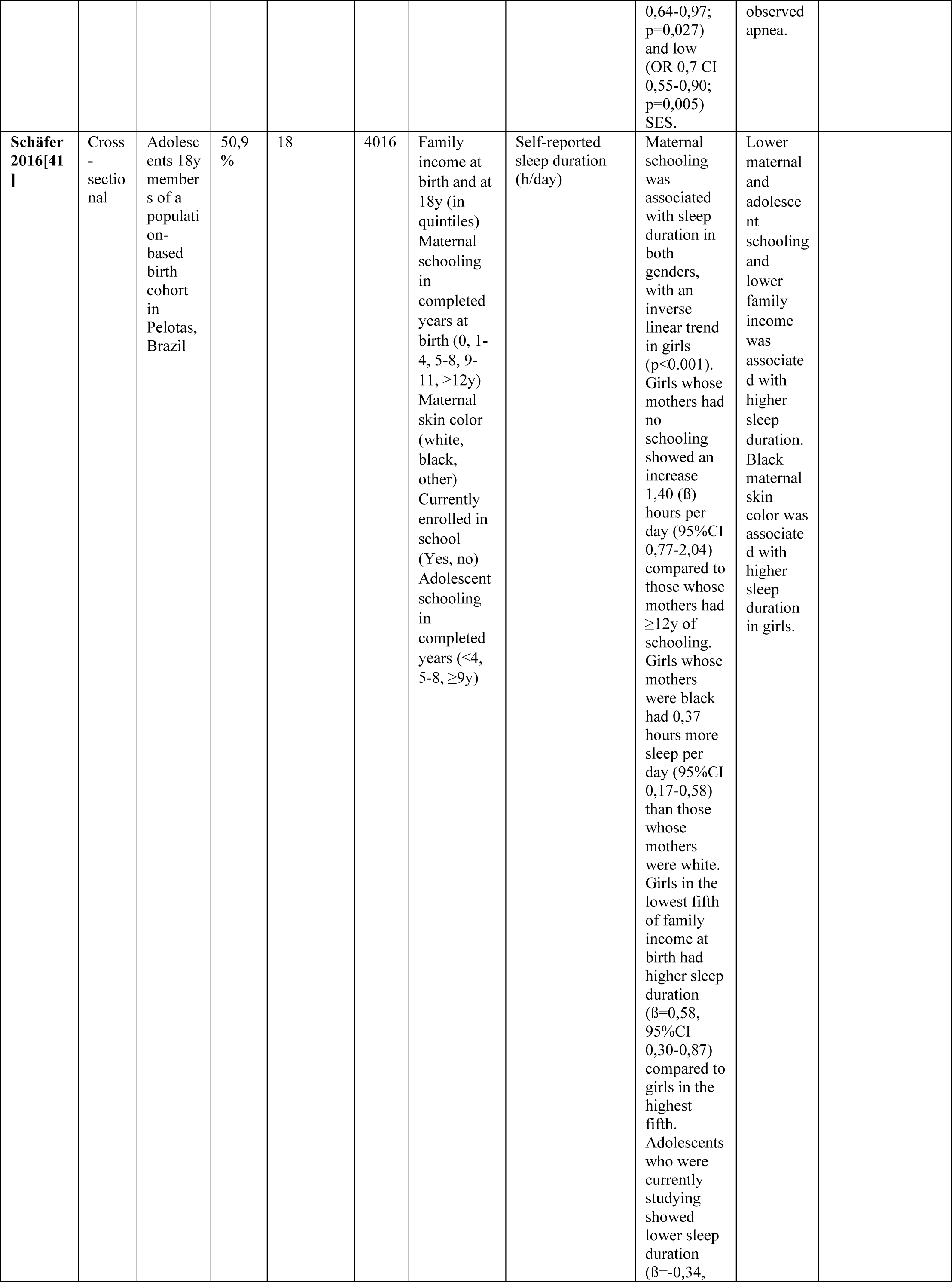
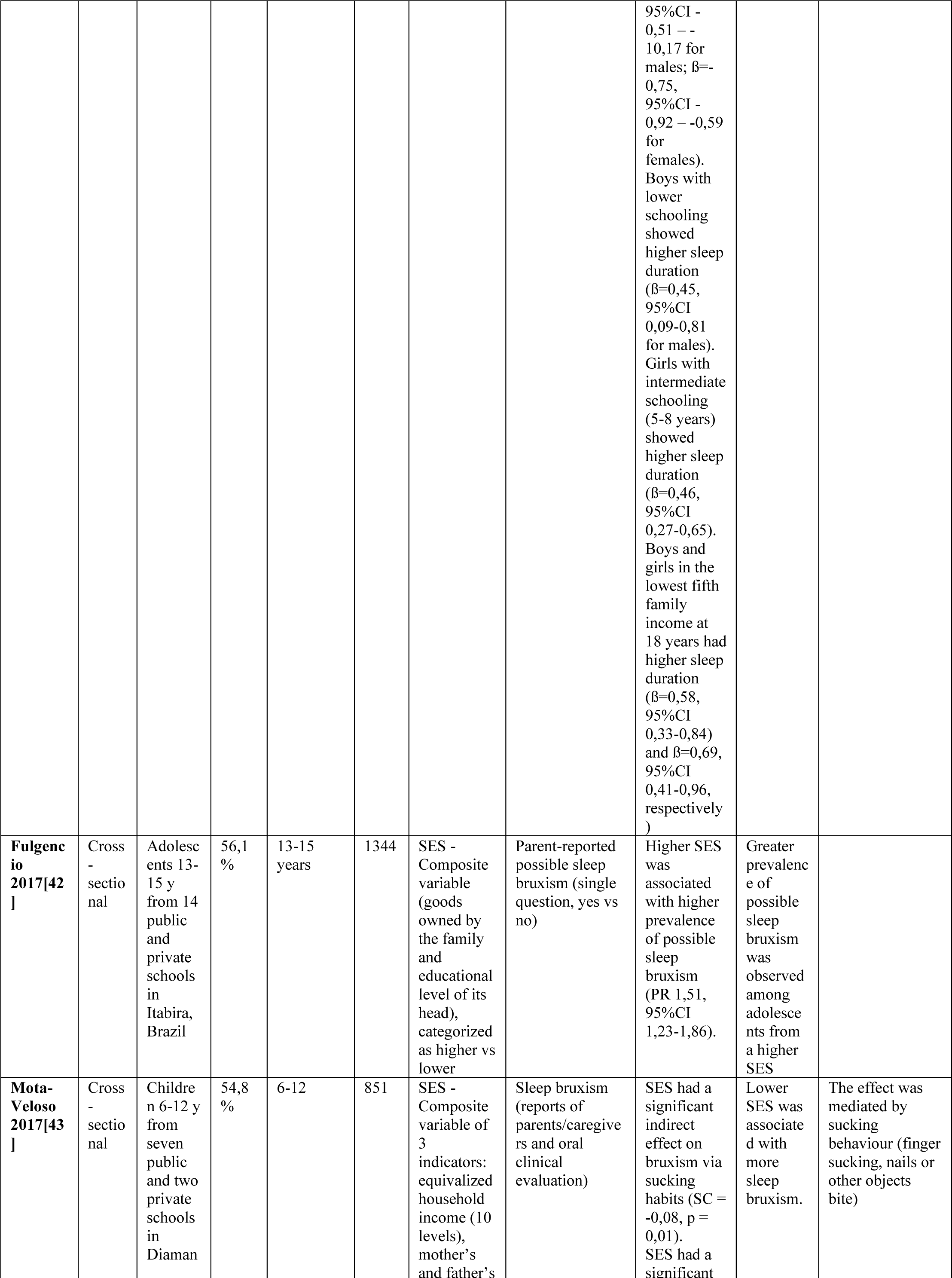
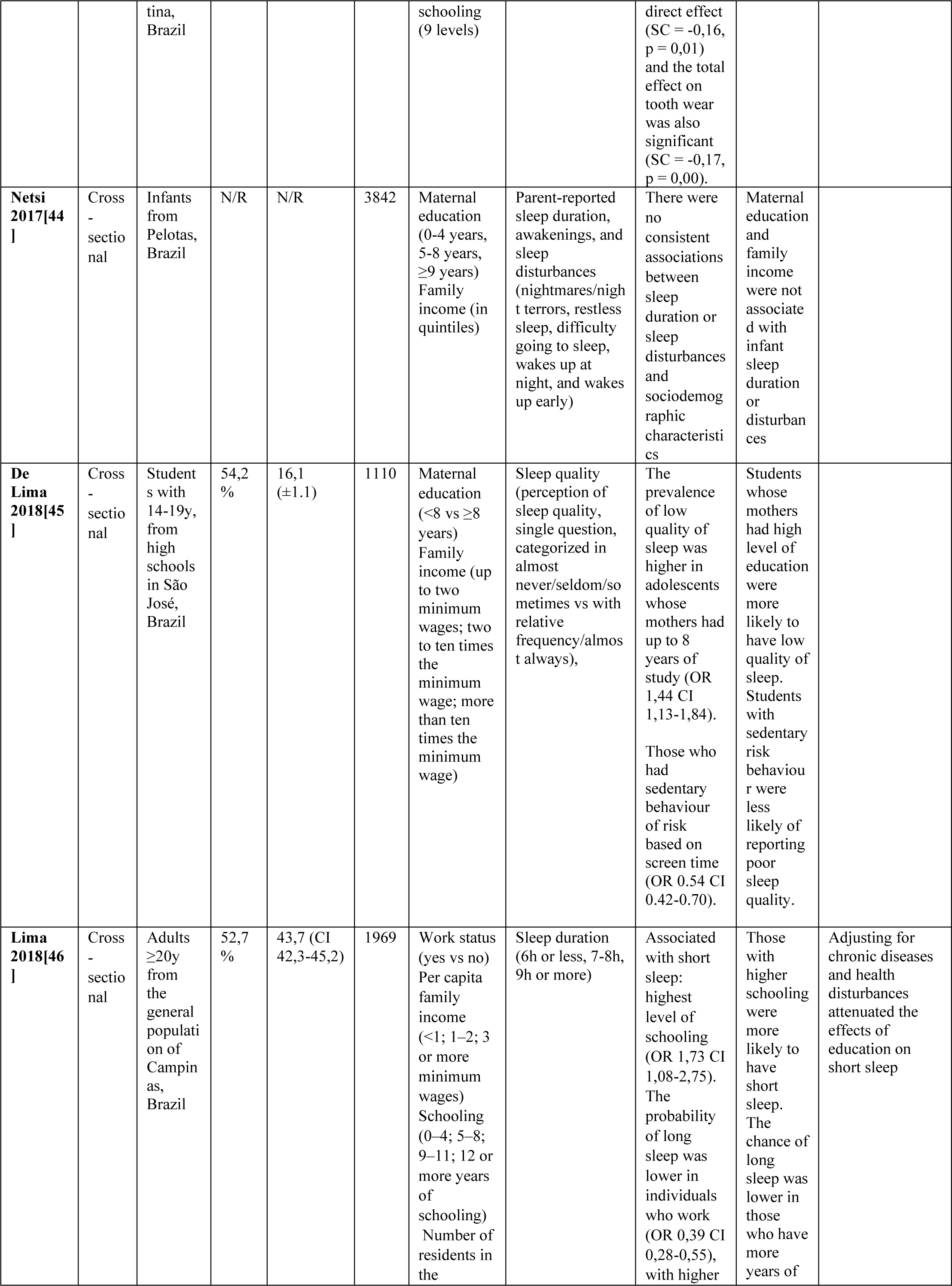
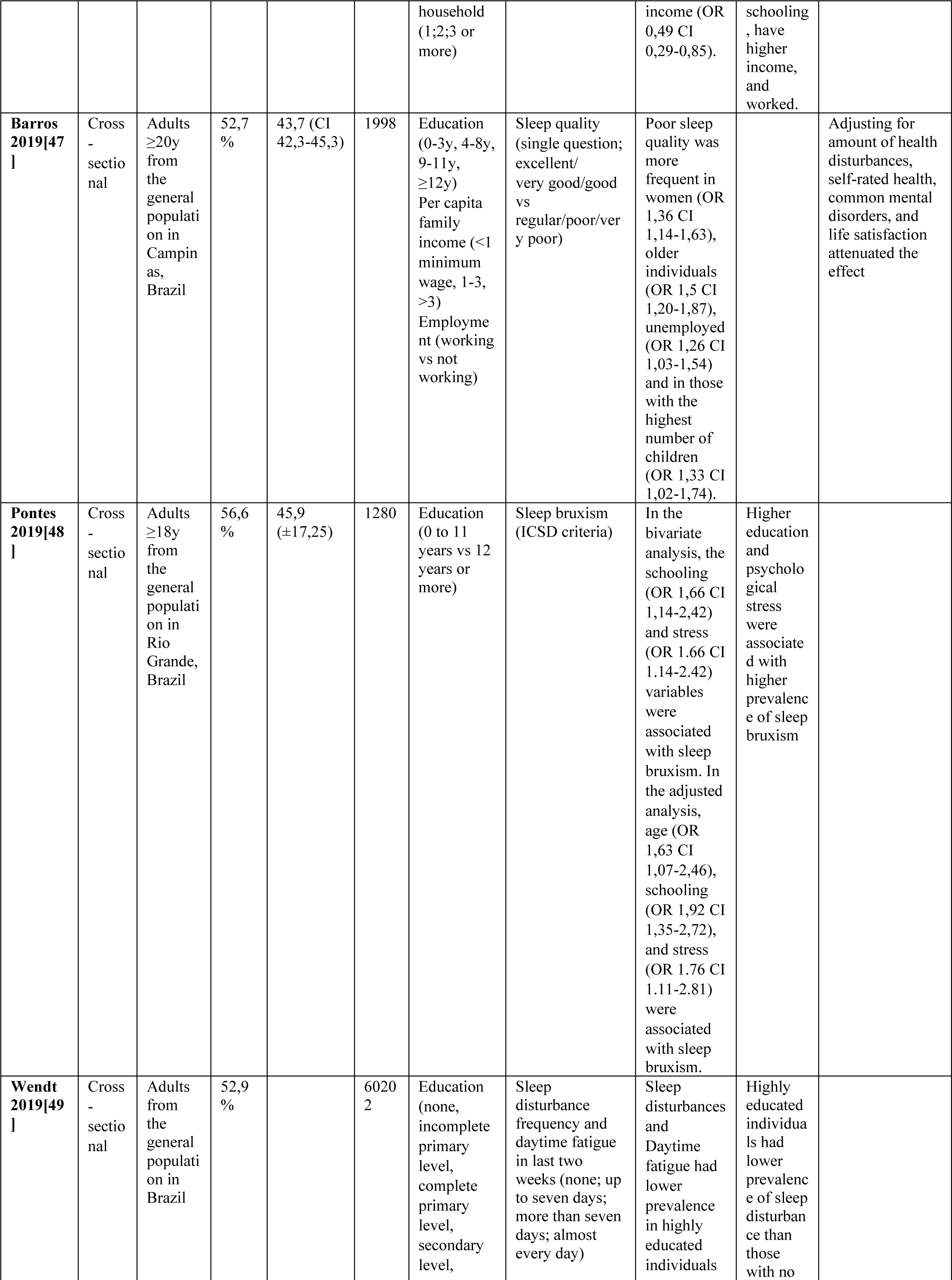
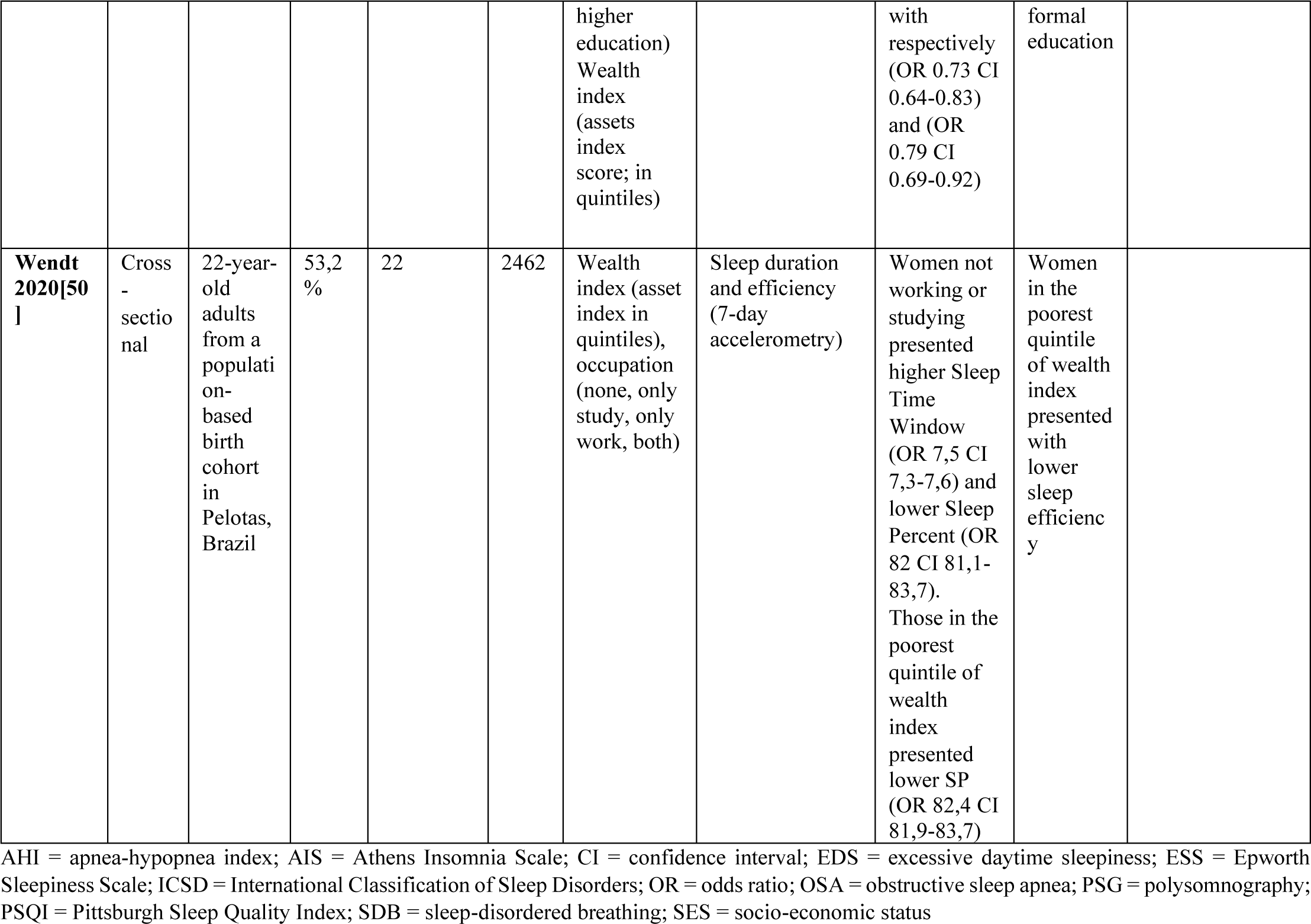
Characteristics of included studies investigating determinants of sleep health in Latin America.

**Table 2:** Quality rating of the included studies using the NIH quality assessment tool.

| Study | Q1 | Q2 | Q3 | Q4 | Q5 | Q6 | Q7 | Q8 | Q9 | Q10 | Q11 | Q12 | Q13 | Q14 | Quality rating |
| --- | --- | --- | --- | --- | --- | --- | --- | --- | --- | --- | --- | --- | --- | --- | --- |
| Rocha 2002 [31] | Y | Y | Y | Y | Y | N | N | Y | N | N | Y | NA | NA | Y | Fair |
| Hara 2004[32] | Y | Y | Y | Y | Y | N | N | Y | N | N | Y | NA | NA | Y | Fair |
| Blay 2008[33] | Y | Y | Y | Y | Y | N | N | Y | Y | N | N | NA | NA | Y | Fair |
| Fritsch Montero 2010[34] | Y | Y | N | Y | N | N | N | Y | Y | N | Y | NA | NA | Y | Fair |
| Tufik 2010[35] | Y | Y | Y | Y | Y | N | N | Y | Y | N | Y | NA | NA | Y | Good |
| Blümel 2012[36] | Y | Y | Y | Y | Y | N | N | Y | Y | N | Y | NA | NA | Y | Good |
| Lima 2012[37] | Y | Y | Y | Y | Y | N | N | Y | N | N | N | NA | NA | Y | Fair |
| Hoefelmann 2013[38] | Y | Y | Y | Y | Y | N | N | Y | N | Y | Y | NA | NR | Y | Good |
| Carrillo- Larco 2014[39] | Y | Y | Y | Y | N | N | N | Y | Y | N | Y | NA | NA | Y | Fair |
| Schwartz 2015[40] | Y | Y | Y | Y | N | N | N | Y | Y | N | Y | NA | NA | Y | Fair |
| Schäfer 2016[41] | Y | Y | Y | Y | N | N | N | Y | Y | Y | N | N | Y | Y | Good |
| Fulgencio 2017[42] | Y | Y | Y | Y | Y | N | N | Y | Y | N | Y | NA | NA | Y | Good |
| Mota-Veloso 2017[43] | Y | Y | Y | Y | Y | N | N | Y | Y | N | Y | NA | NA | Y | Good |
| Netsi 2017[44] | Y | Y | Y | Y | N | N | N | Y | N | Y | N | N | Y | Y | Fair |
| De Lima 2018[45] | Y | Y | Y | Y | Y | N | N | Y | Y | N | Y | NA | NA | Y | Good |
| Lima 2018[46] | Y | Y | Y | Y | Y | N | N | Y | Y | N | N | NA | NA | Y | Fair |
| Barros 2019[47] | Y | Y | Y | Y | Y | N | N | Y | Y | N | N | NA | NA | Y | Fair |
| Pontes 2019[48] | Y | Y | Y | Y | Y | N | N | N | Y | N | Y | NA | NA | Y | Fair |
| Wendt 2019[49] | Y | Y | Y | Y | Y | N | N | N | N | N | N | NA | NA | Y | Poor |
| Wendt 2020[50] | Y | Y | Y | Y | Y | N | N | Y | Y | N | Y | NA | N | Y | Good |

\*N: No; NA: not applicable; NR: not reported; Y: Yes

### 3.2 Descriptive synthesis of articles

Quantitative analyses are presented in the results section. Presented below is a descriptive analysis, which provides a deeper examination of the overall findings that were not considered in the quantitative analysis. Table 1 presents details of the individual studies included in the descriptive analysis.

#### Sleep duration

Seven cross-sectional studies examined the relation between socioeconomic status (SES) and sleep duration [37–39, 41, 44, 46, 50]. One of these was conducted with children (from 3 to 48 months old) [44], two studied adolescents (12 to 19 years old) [38, 41], three involved adults [37, 46, 50] and one encompassed both adolescents and adults [39].

Overall, a higher SES was associated with shorter sleep duration. Specifically, highest level of education [37–39, 41, 46], being employed [37–39, 46], higher income [38, 39, 41, 46] and living with more residents [46] were associated with shorter sleep duration in samples of adults and adolescents. In addition, long sleep was more prevalent among housewives [37], adolescents with black maternal skin color [41] and whose mothers had lower schooling [41].

In contrast, the poorest wealth index and being unemployed or not studying were associated with lower sleep percentage in another study with adults[50]. One study did not find consistent associations between sleep duration and maternal education or family income in children [44]. The overall quality of the selected studies was good for three studies [38, 41, 41] and fair for four studies [37, 39, 44].

#### Sleep quality/sleep disturbance

Eight cross-sectional studies assessed the relationship between sleep quality or disturbance and SES [33, 34, 36, 38, 44, 45, 47, 49]. One of these studies was conducted with infants (from 3 to 48 months) [44] two with students (14 to 19 years old) [38, 45] and the others with adults [33, 33, 47, 49]. One study focused only on adult women [36]. The overall quality of the studies selected was good for three studies [36, 38, 45], fair for three studies [33, 34, 47] and poor for one study [49].

Globally, lower SES was associated with diminished sleep quality. Specifically, low income [33, 38] and unemployment [34, 47] were associated with impaired sleep quality. Two studies indicated a higher prevalence of sleep disturbance in women [34, 47]. More educated adults had significantly less sleep disturbance [33, 36, 49]. In contrast, one study found that higher maternal education was associated with lower quality of sleep in students [45], but this association was not found in infants in another study [44]. Additionally, psychiatric comorbidities [33, 34], alcohol and drug consumption [34, 36] were also associated with sleep disturbance.

#### Insomnia

Concerning insomnia, two cross-sectional studies assessed its relationship with SES [31, 36]. One study evaluated only women [36] and one study assessed adults in general[31]. The overall quality of the two studies was good.

In both of the aforementioned studies, insomnia was independently associated with individuals with less education [31, 36]. Moreover, alcohol and drug consumption was also associated with insomnia, according to another study [36].

#### Excessive daytime sleepiness

Three cross-sectional studies approached the association between excessive daytime sleepiness and SES [32, 40, 49]. All studies assessed adults. The overall quality of the studies selected was good for one study [32], fair for another study [40] and poor for the third study [49].

Largely, excessive daytime sleepiness was associated with lower SES in one the studies [40].Additionally, it was also significantly more prevalent in individuals with lower family income [32] and less education [49].

#### Obstructive sleep apnea (OSA) / Sleep-disordered breathing (SDB) symptoms

Concerning OSA/SDB, two cross-sectional studies assessed their relationship with SES [35, 40]. Both studies evaluated adults in the general population. The overall quality was good in one study [35] and fair for the other study [40].

In one study, lower SES was associated with less frequent snoring. However, no significant association was found between SES and observed apneas [35]. The other study did not find any association between SES and OSA [40].

#### Bruxism

Three cross-sectional studies assessed the relation between SES and bruxism [42, 43, 48]. One of the studies was conducted with infants (ages 6-12 years)[43], one with adolescents (aged 13-15 years) [42] and the last one with adults [48]. The overall quality was good in two studies [42, 43] and fair in one study [48].

Among these three studies, two studies reported that sleep bruxism was independently associated with higher SES, including higher education[42, 48]. However, the other study, conducted by Mota-Veloso *et al*. [43], found out that SES had a significant indirect effect on bruxism via sucking habits (finger sucking, nails or other objects bite) and that lower SES was associated with more sleep bruxism [43].

### 3.3 Prevalence of sleep disturbances in Latin America

Based on our selection criteria, eighteen studies [31–40, 42–49] were eligible. Where results in the same article were reported for different sleep disturbance types separately, they were entered into the analysis as separate studies. Therefore, a total of 18 papers including 28 studies were included in the final meta-analysis which is shown in **Fig 2.A**.

**Fig 2:**
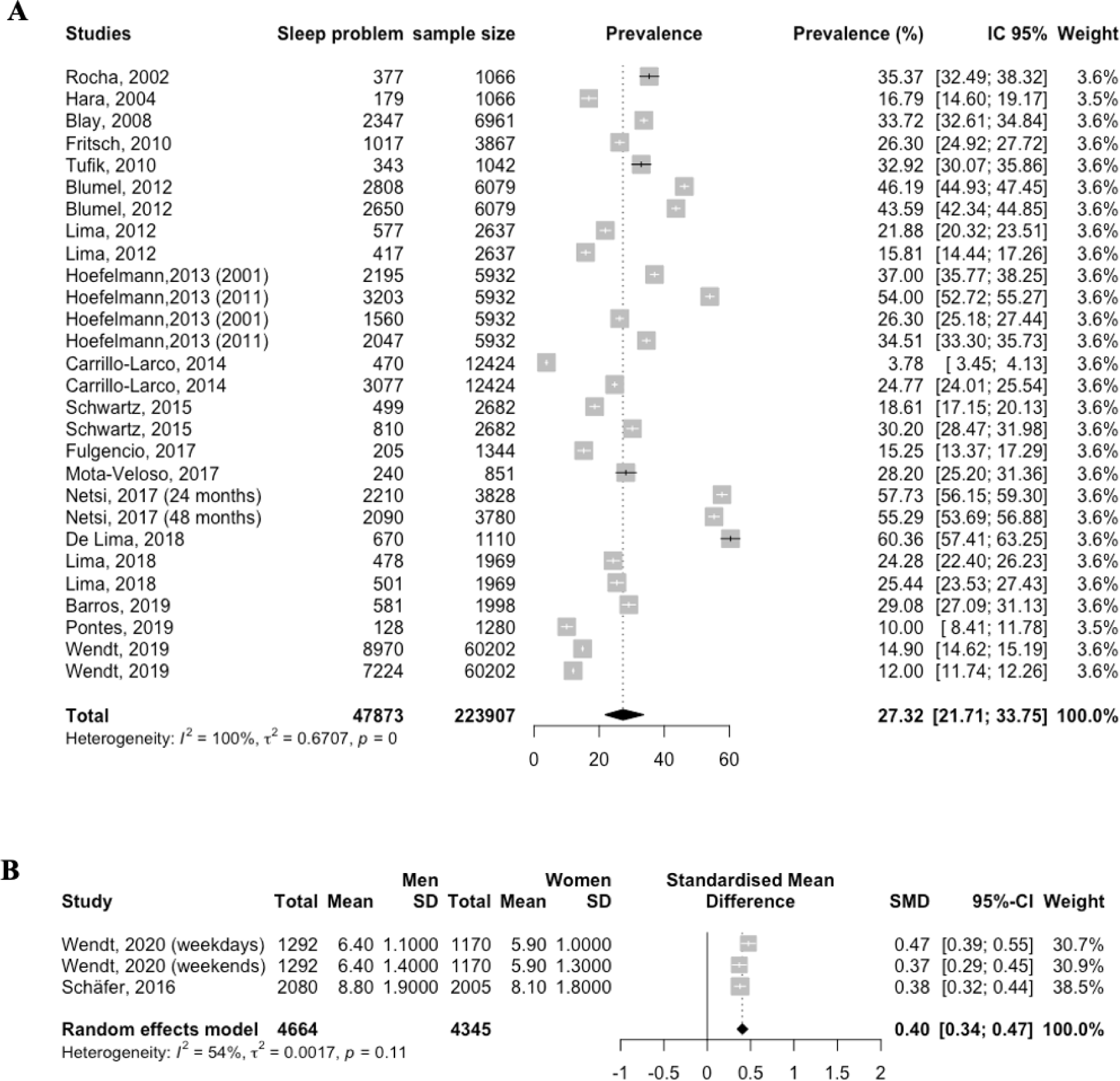
Forest plot showing the primary outcomes in 31 cross sectional studies from 20 published reports in Latin America: **(A)** Prevalence of sleep disturbances; **(B)** Sleep length (hours)

The overall pooled prevalence for sleep disturbance in Latin America was 27.32 % (95 % CI 21.71–33.75, I^2^ = 100 %) (**Fig 2.A**). To decide whether to include all of the articles examining sleep disturbances or not, a publication bias chart was created. The results showed that publication bias was not significant (*p* = 0.059) (**Fig 3**).

**Fig 3:**
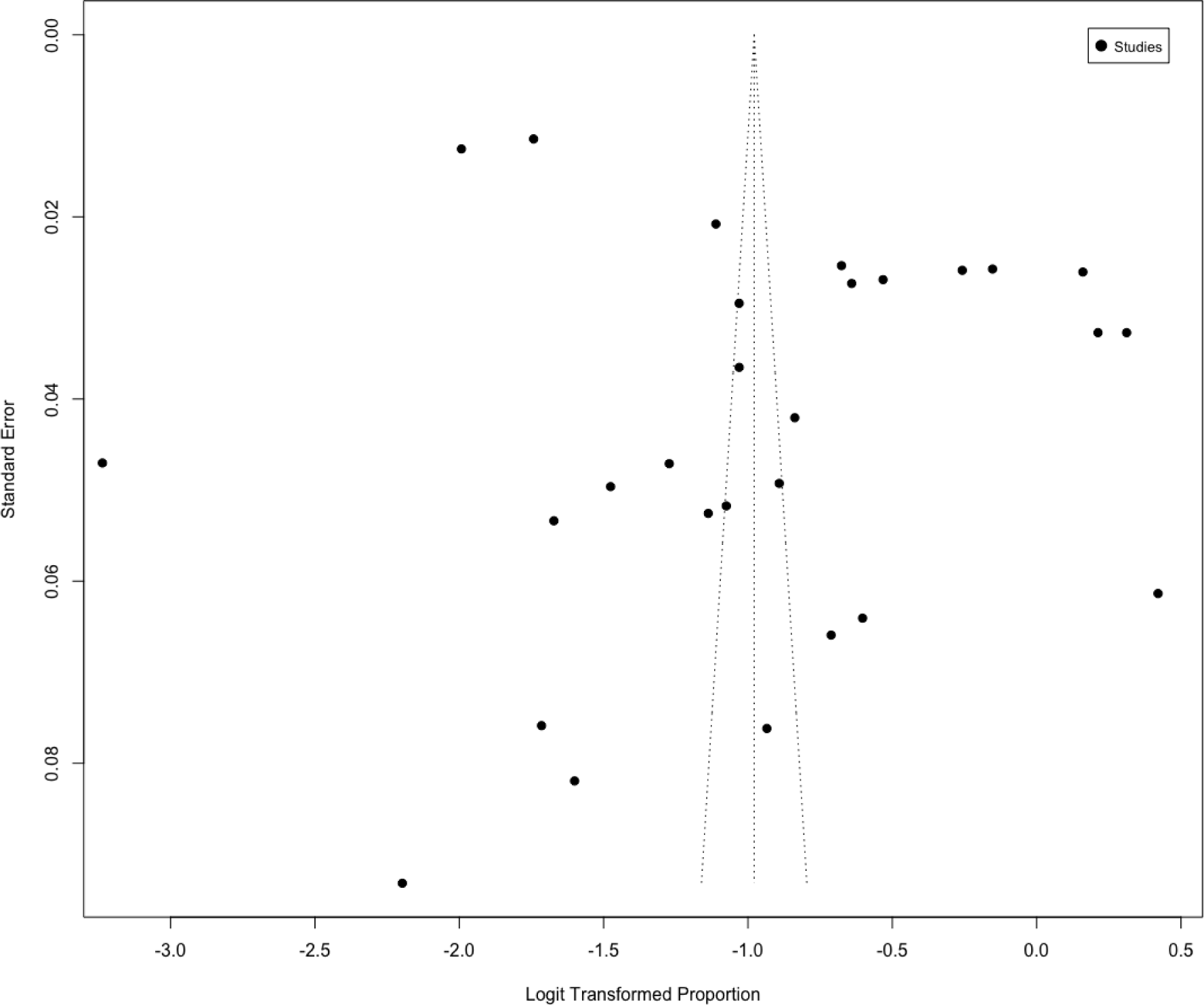
Funnel plot for meta-analysis of the prevalence of sleep disturbances in Latin America. Egger’s test: .0598.

We divided eighteen papers into seven categories according to different types of sleep disturbances. Subgroup analysis was manipulated based on seven categories (**Fig 4**). The highest prevalence was for insomnia, with 39.52 % (95 % CI 31.79–47.82; *p* < 0.01), and the lowest prevalence was for EDS, with 15.52 % (95 % CI 11.85–20.07; *p* < 0.01). The overall pooled prevalence for sleep disturbance was 37.22 % (95 % CI 27.73– 47.82; *p* = 0) across eight articles [33, 34, 36, 38, 44, 45, 47, 49], for OSA/SDB was 31.32 % (95 % CI 28.77–34.00; *p* = 0.11) across two articles [35, 40] and for bruxism was 16.60 % (95 % CI 8.83–29.04; *p* < 0.01) across three articles [42, 43, 48]. The overall pooled prevalence for short sleep duration was 23.09 % (95 % CI 8.98– 47.75; *p* =0) across four articles [37–39, 46], and for long sleep duration was 21.67 % (95 % CI 15.95–28.75; *p* < 0.01) across three articles [37, 39, 46].

**Fig 4:**
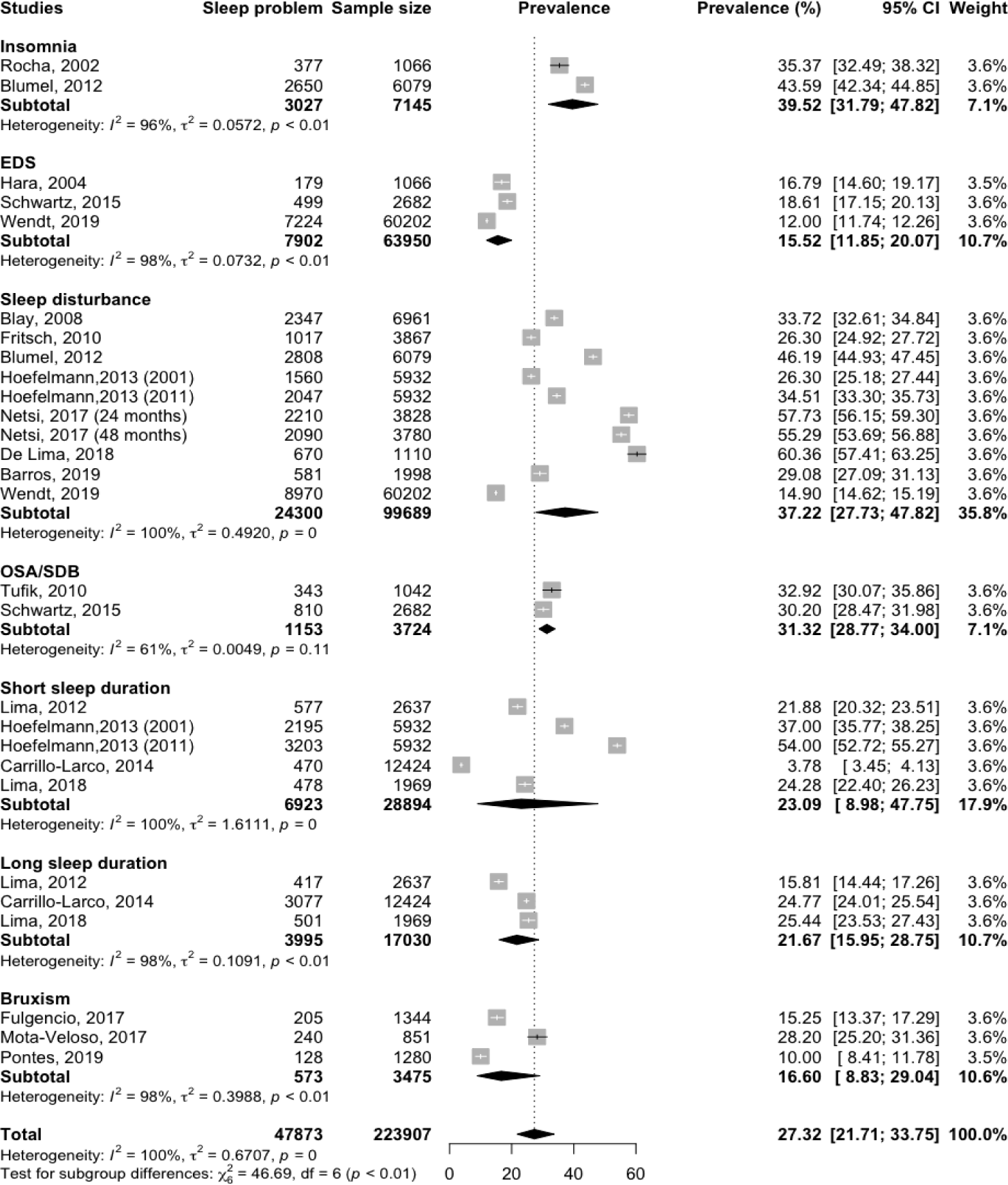
Subgroup analysis on the prevalence of sleep disturbances in Latin America by type of sleep disturbance. The black dot point is the estimate and the horizontal line is the 95% CI for prevalence plotted for each study. The black diamond at the bottom of each type of sleep disturbance is the estimates average prevalence. CI: confidence interval

### 3.4 Sleep length in Latin America

A total of two articles [41, 50] reported the sleep length (**fig 2.B**). Where results were reported for men and women separately, they were entered into the analysis as separate studies. In the pooled analysis, sleep length was significantly higher in men with sleep disturbances than in women with sleep disturbances with a standardized mean of 0.40 hours (95% CI: 0.34-0.47; *p* = 0.11; I^2^ = 100%).

### 3.5 Subgroup analysis

Because of significant heterogeneity across the included studies, a subgroup analysis was performed by region (cities), age groups and study quality, in relation to principal outcome variable. The analysis revealed that the prevalence of sleep disturbances was highest in Brazil 28.52 (95 % CI 22.38-35.57) than Peru 15.91 (95 % CI 6.17-35.27). Also, the pooled prevalence of sleep disturbances among infant 56.52 (95 % CI 54.11-58.89) was greater than adolescents 27.11 (95 % CI 16.55-41.09) and adults 24.24 (95 % CI 18.93-30.48) (**Table 3**).

**Table 3:** Subgroup analyses for the prevalence of sleep disturbances in Latin America.

| Subgroup | Number of studies | Pooled Prevalence (95 CI%) | I<sup>2</sup> (%) |
| --- | --- | --- | --- |
| **Total** | 28 | 27.32 [21.71-33.75] | 100 |
| **Cities** |  |  |  |
| Brazil | 21 | 28.52 [22.38-35.57] | 100 |
| Chile | 1 | - | - |
| Peru | 4 | 15.91 [6.17-35.27] | 100 |
| Multicentric | 2 | 44.89 [42.36-47.45] | 88 |
| **Age group** |  |  |  |
| Adult | 15 | 24.24 [18.93-30.48] | 100 |
| Adolescents | 10 | 27.11 [16.55-41.09] | 100 |
| Children | 1 | - | - |
| Infant | 2 | 56.52 [54.11-58.89] | 78 |
| **Study's quality** |  |  |  |
| Good | 12 | 34.73 [27.07-43.28] | 99 |
| Fair | 14 | 24.08 [16.61-33.55] | 100 |
| Poor | 2 | 13.38 [10.79-16.49] | 100 |
| Infant | 2 | 56.52 [54.11-58.89] | 78 |

### 3.6 Risk factors

Meta-analysis was possible for sleep disturbances prevalence with three SES factors (Education, income and employment status).

#### 3.6.1 Education and Sleep disturbances

Eight studies [31, 33, 36, 37, 39, 45, 48, 49] evaluated the association between education and prevalence of sleep disturbances as shown in **Fig 5.A**. There was found no association between low education and prevalent sleep disturbances, with a pooled OR of 1.42 (95%CI [0.87-2.29]; *p*=<0.01). However, the meta-analysis showed that the high educated population had a lower prevalence of sleep disturbances (OR 0.83; 95%CI [0.69-0.99]; *p*=<0.01), with a high heterogeneity between studies (I^2^= 79%).

**Fig 5:**
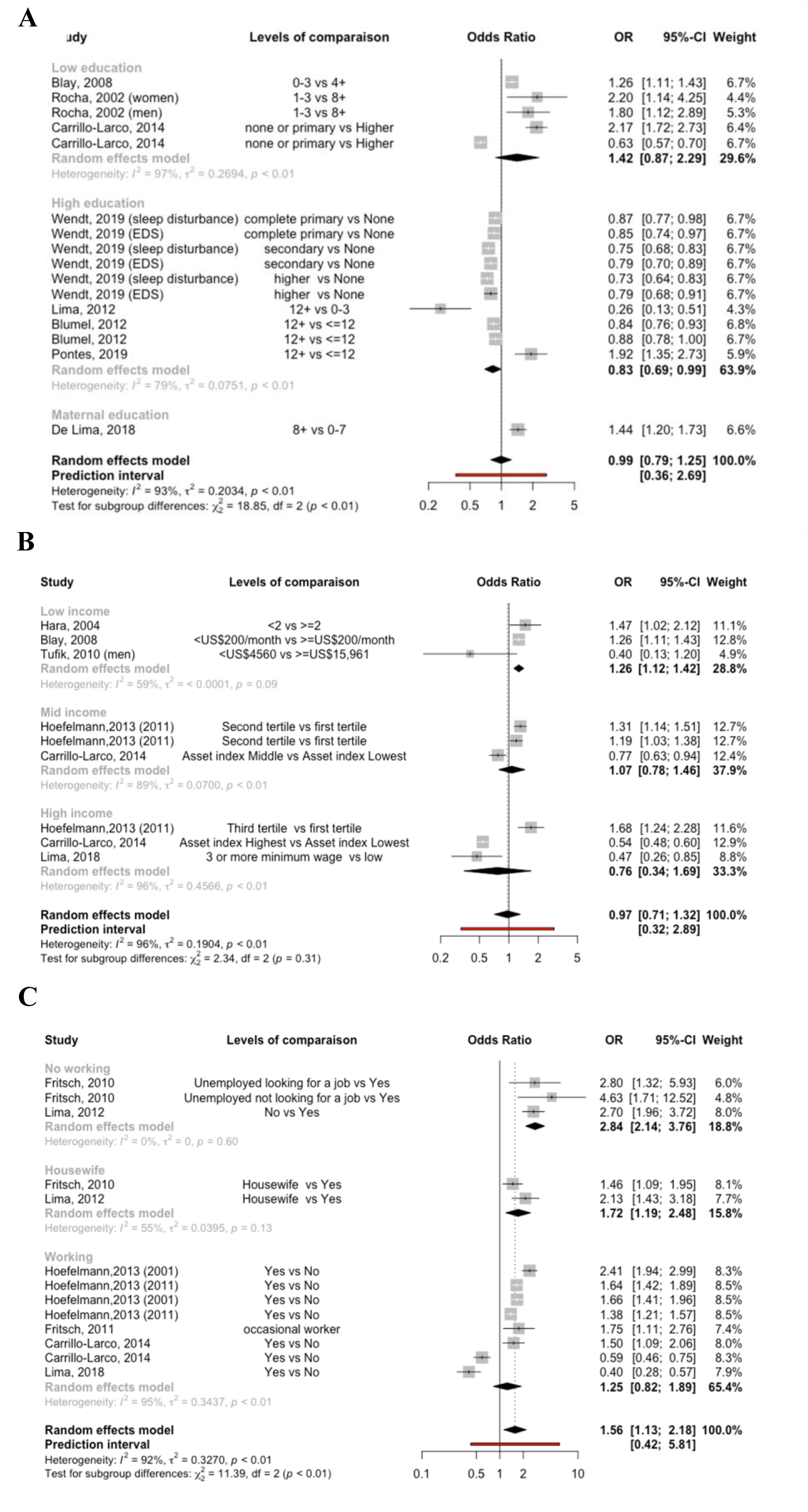
(A) Forest plot for education and sleep disturbances prevalence (compared to the reference group). (B) Forest plot for income and sleep disturbances prevalence (compared to the reference group). (C) Forest plot for employment status and sleep disturbances prevalence (compared to the reference group). **C** Box sizes reflect the weights of studies included in the meta-analysis, horizontal lines are the 95% CIs, and the summary OR are represented by the diamond. OR: odds ratio, CI: confidence interval

Separating the education analyses by quality of studies did not reveal a significant subgroup effect for sleep disturbances prevalence (*p=0.70*; **Fig 6.A**). Similarly, when the education analyses were separated by cities, no significant subgroup effect for the prevalence of sleep disturbances was observed (*p=0.70*; **Fig 6.B**).

**Fig 6:**
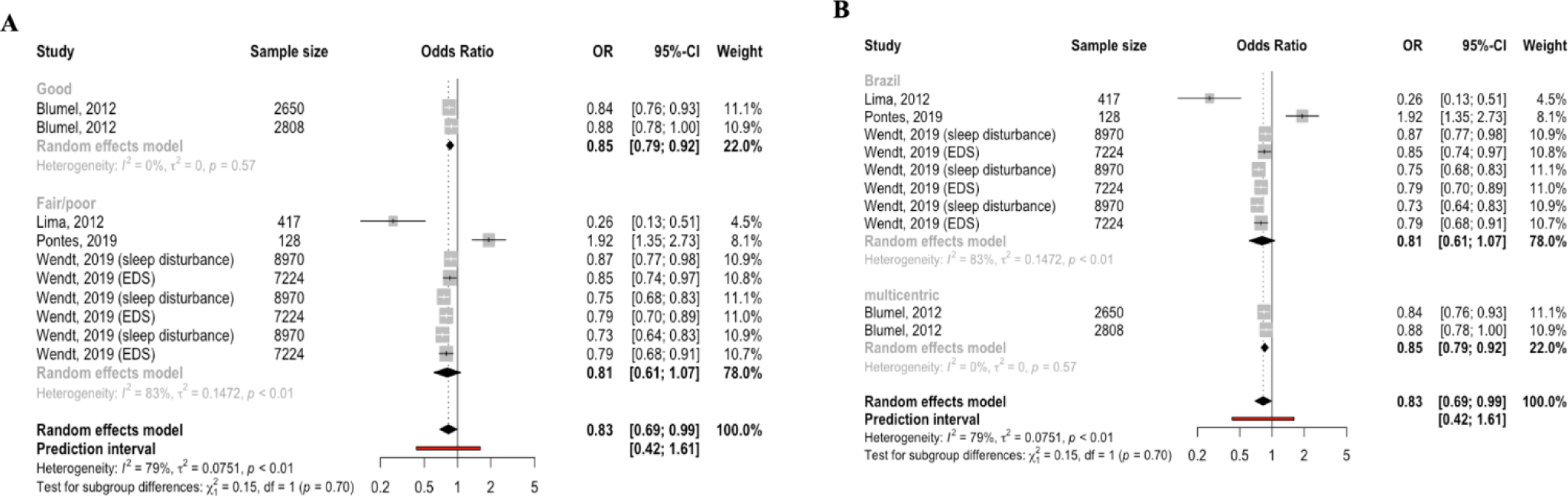
Subgroup analyses to explore sources of heterogeneity in risk factors of sleep disturbances. **(A)** Forest lot demonstrating that higher education was associated with sleep disturbances prevalence by quality of the study good vs fair/poor)**. (B)** Forest plot demonstrating that higher education was associated with sleep disturbances revalence by City (Brazil vs multicentric)

**Fig. 7:**
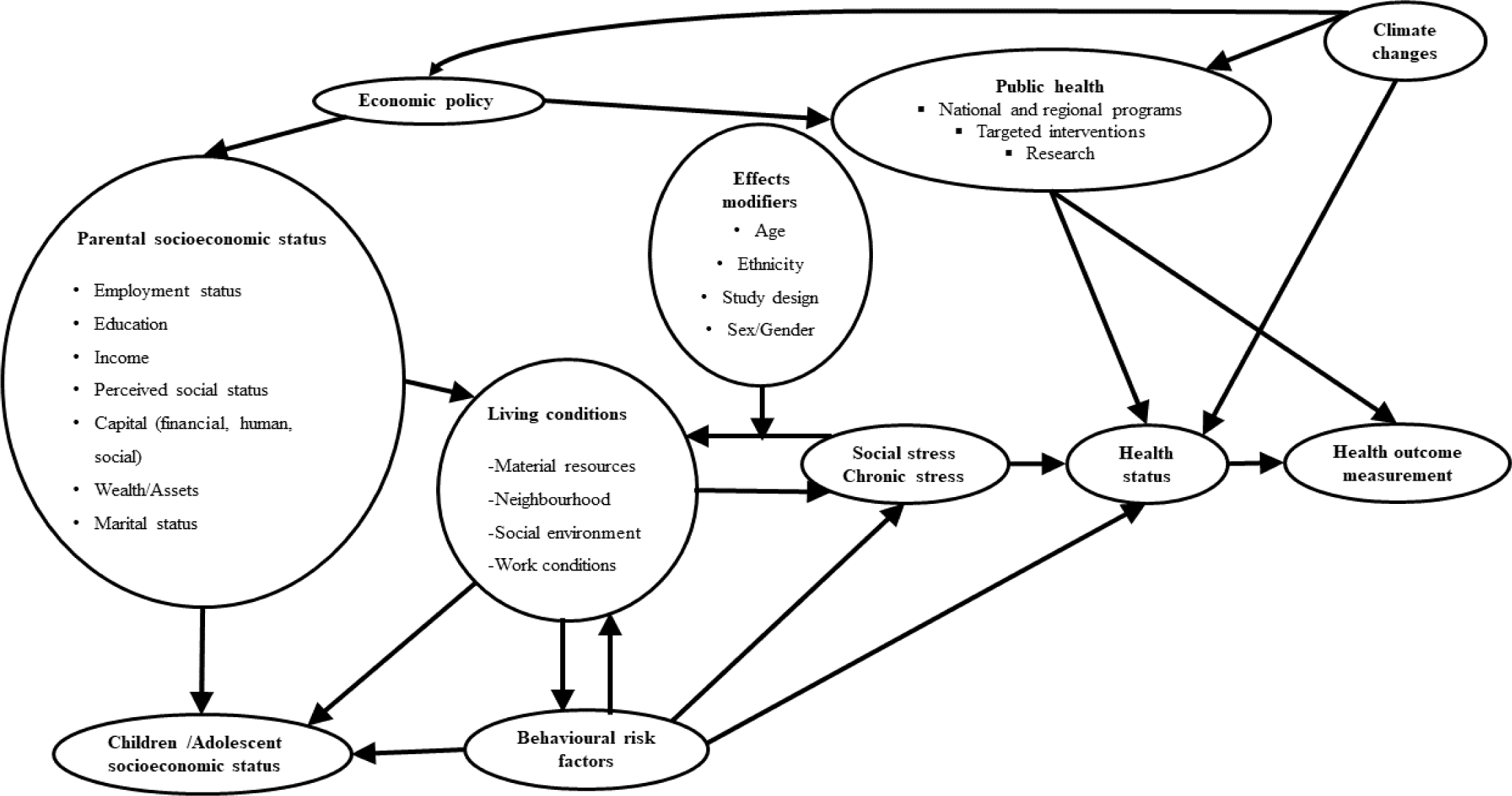
Ecoepidemiological model of sleep disturbances

#### 3.6.2 Income and Sleep disturbances

Six studies [32, 33, 35, 38, 39, 46] considered income as a risk factor of sleep disturbances in Latin America. The meta-analysis revealed a significant relationship between low income and the prevalence of sleep disturbances (OR 1.26; 95%CI [1.12-1.42]; *p*=0.09), with moderate heterogeneity between studies (I^2^= 59%). However, it did not show a significant association between middle income (OR 1.07; 95%CI [0.78-1.46]; *p*=<0.01) or high income (OR 0.76; 95%CI [0.34-1.32]; *p*=<0.01) and prevalence of sleep disturbances (**Fig 5.B**).

#### 3.6.3 Work and Sleep disturbances

Data on the relationship between work and sleep disturbances are shown in **Fig 5.C.** The meta-analysis showed a significant relationship between no working and the prevalence of sleep disturbances (OR 2.84; 95%CI [2.14-3.76]; *p*=0.09) with no heterogeneity between studies (I^2^= 0%). Similarly, in the pooled analysis, being housewife was associated with high prevalence of sleep disturbances (OR 1.72; 95%CI [1.19-2.48]; *p*=0.13), exhibiting moderate heterogeneity across studies (I^2^= 55%).

## 4 DISCUSSION

### 4.1 Detailed summary of findings

The most used determinants were income (monthly personal income, monthly family income, per capita income, annual household income), wealth/assets, number of residents in the household, employment status/occupation, accessed healthcare system and composite indices. The sleep outcomes analyzed were excessive daytime sleepiness (EDS), sleep duration, sleep quality/sleep disturbance, insomnia, obstructive sleep apnea (OSA)/sleep-disordered breathing (SDB) symptoms and bruxism. Higher SES was associated with lower sleep duration. Lower SES was associated with a decrease in sleep quality. EDS was significantly more prevalent in individuals with lower family income and less education. Sleep bruxism was associated with higher education and lower SES was associated with more sleep bruxism. Among the 20 included articles, 12 were rated as fair or poor in study quality. Therefore, a meta-analysis was performed to estimate the prevalence of sleep disturbances in Latin America and the main SES risk factors that could be associated with it. The pooled prevalence using a meta-analysis of the random effects model was 27.32 % (95 % CI 21.71–33.75), with high heterogeneity (I^2^ = 100 %). The meta-analysis showed that the prevalence of sleep disturbances decreased with high education (OR 0.83; 95%CI [0.75-0.91]; I^2^= 79%), while it increased with low income (OR 1.26; 95%CI [1.12-1.42]; I^2^= 59%), unemployment (OR 2.84; 95%CI [2.14-3.76]; I^2^= 0%) and being housewife (OR 1.72; 95%CI [1.19-2.48]; I^2^=55.4%).

### 4.2 Relationship with public health literature

Epidemiologic data continues to increase the literature about the influence of sleep on general population’s health status. Sleep plays a vital role in several body functions as well as health disparities. Nowadays, scientific community is still investigating external and environmental factors affecting sleep mechanisms, but there are still a lot unknown. Based on current findings, it can be hypothesized that sleep disturbances are associated with socioeconomic status liked suggested many other studies [17, 51–58]. The fact is, gradient of health disparity existing for some diseases like cardiovascular illness, seems the same for sleep. Regardless of world region where investigation is made, sleep disparities are observed because our findings on Latin America supports previous results [17, 51–58]. Our findings are coherent with previous studies elsewhere than Latin America and they are additional arguments in favor of the establishment of a more efficient worldwide program framing sleep health management.

### 4.3 The necessity of a multidimensional sleep management

The management of sleep disturbances should be addressed through a multidimensional approach. The majority of studies used to focus on biological and psychological factors influencing sleep while it was only recently that SES was recognized as major determinant of sleep. Recent epidemiological studies performed outside Latin America in different public health contexts, associated sleep with stress [4], work conditions [8], environment [59], employment [17, 60, 61] and revealed latent interactions existing between government policy and public health strategies [22, 55, 62–64]. Obviously, a government’s economic policy influences funding of public health programs. Similarly, SES influences directly health status regardless disease assessed through individual’s living conditions and their resulting behavioral risk factors and stress. Knowing that sleep disparities can be measured objectively and quantitatively [6, 7, 50], our suggestion for governments is to invest as soon as possible in preventive management programs of sleep disturbances; before they become uncontrollable. It was already documented how expensive for economy were absenteeism and presenteeism due to sleep disturbances [65, 66], but diverse governments didn’t move forward yet with strong regulations to reduce these important losses [67, 68].

In this meta-analysis, there is an unequal distribution of research, with 80% of studies originating from Brazil (**Table 1**). Even if Brazil is representative of Latin American populations, its public health’s context regarding sleep management is not necessarily identical for its neighbors. More research should be performed in other Latin American countries to obtain an accurate overview of sleep disparities in this continent. Our suggestion for scientists, is to not forget that cross-sectionals studies are often used to understand determinants of health and establish preliminary evidence [69]; but are useless when it is necessary to consider correlation between theoretical determinant and health outcome. This first meta-analysis on sleep determinants in Latin America, enlighten the high quality of cross-sectional studies published but also the lack of systematic review and longitudinal studies. To support public health strategies, randomized controlled trial and longitudinal studies are required [69].

## Data Availability

All data used for this review are available publicly and already published.

## Acknowledgements

None

## Funding

This research did not receive any specific grant from funding agencies in the public, commercial, or not-for-profit sectors.

## Author contribution

*FAES:* Conceptualization, Methodology, Validation, Investigation, Data Extraction, Writing - Original and Revised Draft, Review and Editing. *FTS:* Data Extraction, Writing– Original Draft. *RQR:* Data Extraction, Writing– Original Draft. *MC:* Data Extraction, Writing – Original Draft. *SZ:* Methodology, Data Analysis, Meta-Analysis, Writing - Review and Editing. *GCZ:* Writing– Original Draft and Editing.

## Conflicts of interest

The authors declare they have nothing to disclose. If only some authors declare conflicts of interest, detail those conflicts and then include the following statement: All other authors declare they have nothing to disclose.

## Notes

### Competing Interest Statement

The authors have declared no competing interest.

### Funding Statement

This study did not receive any funding

### Summary of Updates

Figures 2 and 3 has been removed and replace by a new theoretical model explaining a different approach. All pages orientation were changed to portrait as requested by the medRxiv team

